# Understanding and Addressing Vaccine Hesitancy in the Context of COVID-19: Development of a Digital intervention

**DOI:** 10.1101/2021.03.24.21254124

**Authors:** Holly Knight, Ru Jia, Kieran Ayling, Katherine Bradbury, Katherine Baker, Trudie Chalder, Joanne R Morling, Lindy Durrant, Tony Avery, Jonathan Ball, Caroline Barker, Robert Bennett, Tricia McKeever, Kavita Vedhara

## Abstract

**Background:** Severe Acute Respiratory Coronavirus 2 (SARS-CoV-2) was identified in late 2019, spreading to over 200 countries and resulting in almost two million deaths worldwide. The emergence of safe and effective vaccines provides a route out of the pandemic, with vaccination uptake of 75-90% needed to achieve population protection. Vaccine hesitancy is problematic for vaccine rollout; global reports suggest only 73% of the population may agree to being vaccinated. As a result, there is an urgent need to develop equitable and accessible interventions to address vaccine hesitancy at the population level.

**Method:** We report the development of a scalable digital intervention seeking to address COVID-19 vaccine hesitancy and enhance uptake of COVID-19 vaccines. Guided by motivational interviewing (MI) principles, the intervention includes a series of therapeutic dialogues addressing 10 key concerns of vaccine hesitant individuals. Development of the intervention occurred linearly across four stages. During stage 1, we identified common reasons for COVID-19 vaccine hesitancy through analysis of existing survey data, a rapid systematic literature review, and public engagement workshops. Stage 2 comprised qualitative interviews with medical, immunological, and public health experts. Rapid content and thematic analysis of the data provided evidence-based responses to common vaccine concerns. Stage 3 involved the development of therapeutic dialogues through workshops with psychological and digital behaviour change experts. Dialogues were developed to address concerns using MI principles, including embracing resistance and supporting self-efficacy.

Finally, stage 4 involved digitisation of the dialogues and pilot testing with members of the public.

**Discussion:** The digital intervention provides an evidence-based approach to addressing vaccine hesitancy through MI principles. The dialogues are user-selected, allowing exploration of relevant issues associated with hesitancy in a non-judgmental context. The text-based content and digital format allow for rapid modification to changing information and scalability for wider dissemination.

## Background

Severe Acute Respiratory Coronavirus 2 (SARS-CoV-2) was identified in late 2019. At the time of writing, the latest estimates suggest that it has spread to over 200 countries and has resulted in the deaths of almost two million people.[1] The resulting global pandemic has seriously affected the social and economic fabric of societies everywhere and the physical and mental health crisis continues.[2] Safe and effective vaccines provide a route out of this crisis, but the development of these vaccines, while necessary, are not sufficient. For vaccines to achieve their full potential, the public also need to be willing to be vaccinated. Recent data suggest this cannot be assumed. A recent international survey indicated that, globally, 73% of the population would agree to being vaccinated.[3] This estimate should be viewed against a backdrop of declines in vaccine intent overall and the fact that it masks large variations in intent between countries, genders and across age and ethnic groups.[3–5] Vaccine hesitancy, defined as a “delay in acceptance or refusal of vaccines despite availability of vaccine services” [6] may significantly impact uptake of COVID-19 vaccines, particularly amongst ethnic minorities, women, and those with less education [7]. If, as has been suggested, 75-90% of a population will need to be vaccinated for community protection to be achieved,[8] then there is an urgent need to develop equitable and accessible interventions to address vaccine hesitancy at the population level.

Attempts to improve vaccine uptake are not new and have focussed traditionally on approaches such as information/education, incentives[9–11] and reminders. However, results from successive reviews suggest that the evidence-base in support of any one approach remains limited.[9–12] Furthermore, much of the work has been conducted in the context of adults making decisions for their dependents, rather than adults making decisions for themselves. The generalisability of these findings to COVID-19 vaccines in adults is, therefore, unclear. Nonetheless, much can be gleaned from the existing evidence: information, while necessary, is unlikely to improve vaccine uptake on its own, and interventions need to engage with individuals’ reasons for hesitancy i.e., their hesitancy cognitions.[13]

We report here on the development of a scalable digital intervention which seeks to address the concerns of individuals who are vaccine hesitant with a view to enhancing the uptake of COVID-19 vaccines. We report the process we followed in developing a digital vaccine hesitancy intervention suitable for adults considering a COVID-19 vaccination. In view of the urgency of the public health need, our approach to intervention development was pragmatic and took advantage of existing data where possible and appropriate.

## Methods and Results

Our development involved four main stages and included involvement of public and patient partners throughout:

Stage 1: In order to understand and identify common reasons for COVID-19 vaccine hesitancy and acceptance we carried out a) an analysis of existing survey data collected during the pandemic, b) a rapid systematic literature review and c) an examination of qualitative findings from a series of public engagement workshops regarding immune challenges and vaccines.

Stage 2: We synthesised evidence from independent experts. This entailed qualitative interviews with experts from a range of relevant disciplines to identify evidence-based responses to the most common vaccine concerns raised by the public identified in stage 1.

Stage 3: We developed ‘therapeutic dialogues’ to address common vaccine hesitancy concerns. These were developed in a workshop bringing together experts in psychological and digital behaviour change interventions.

Stage 4: The digital intervention was developed.

As this was a linear process with each stage informing the next, we present the methods and results from each stage consecutively.

## Stage 1: Understanding and identifying common reasons for hesitancy & acceptance

### 1.1 Analysis of existing survey data

As part of an ongoing longitudinal study[14] into the UK population’s mental and physical health over the course of the pandemic, we collected data regarding COVID-19 vaccination intention between 11^th^-30^th^ November 2020. This included whether participants intended to get the vaccine and a free text response to elaborate on their main reasons for this intention. Free text responses were analysed for common themes and the frequency at which these themes appeared was quantified. Where vaccine hesitancy was indicated, themes were also categorised within the WHO 3Cs model of vaccine hesitancy, which proposes that three main factors influence the decision to accept vaccines: confidence, complacency, and convenience[6] Coding and categorisation was conducted independently by two researchers (RJ, KA) with high levels of initial agreement (91% for reasons associated with vaccine hesitancy and 85% for reasons associated with agreement to vaccination). All discrepancies were resolved by discussion.

A total of n=762 individuals provided data (22% of whom indicated they were hesitant about receiving a COVID-19 vaccination); 93% (n=709) of respondents also provided a free-text response indicating their reasons for vaccine acceptance or hesitancy, of which 96% (n=683) provided sufficient detail for reasons to be categorised into themes. For those who expressed vaccine hesitancy, the most common concerns were found to map on to the WHO 3C category of ‘confidence’ (e.g., concerns related to long term complications, side effects and insufficient testing of the vaccines). The second most common concerns related to ‘complacency’ (e.g., beliefs of low personal risk of COVID-19, beliefs in ability to fight off the infection naturally). Concerns related to the ‘convenience’ category were the least common, and centred on a lack of information about the vaccines and altruism (i.e., other people needing the vaccines more) (see Table 1a). In contrast, in respondents who indicated they would be willing to receive a COVID-19 vaccine, the common reasons given related to ‘self-protection’, followed by ‘hope to end the pandemic/wish for normal life’ and a desire to ‘protect the population or unspecified others and control the virus’ (see Table 1b).

**Table 1a:** Common reasons for vaccine hesitancy and acceptance: survey findings

| WHO 3C category | Themes | Count | Examples of free text responses |
| --- | --- | --- | --- |
| **Confidence** | Concerns about unknown long-term effects | 39 | “It hasn’t been long enough to see if there are any long-term risks”<br>“Uncertainties around long-term effects”<br>“Unknown long term side effects” |
|  | Concerns about side effects | 39 | “I don’t have full information about its side effects”<br>“Undiscovered side effects/uncertainty of the side effects”<br>“Unknown long-term side effects” |
|  | Concerns there is an insufficient testing/evidence base | 37 | “Not sure it has been tested thoroughly”<br>“Unclear rigour of the testing/clinical trial results/statistics etc”<br>“It has not been tested at a scale” |
|  | Concerns the development of the vaccine has been rushed | 27 | “Its development and production has been rushed through”<br>“Feels rushed compared to normal vaccine standards”<br>“I don’t think there has been sufficient time to know fully the effects of it” |
|  | Concerns about the safety of the vaccine (but not explicitly side effects) | 21 | “I’m concerned about its safety”<br>“Would want to be 100% sure it was safe”<br>“Would only take it if I was convinced it was 100% safe” |
|  | Unsure about vaccine effectiveness | 14 | “I would like the research evidence about its effectiveness rate (in different age groups especially 60+)”<br>“Not sure about how effective are they, especially as if you get COVID you can get it again. The vaccines antibodies are not as effective as getting the virus itself”<br>“Would prefer a vaccine that stops transmission, not just stop me<br>showing symptoms” |
|  | Concerns around vaccine interactions/<br>effectiveness with existing conditions | 10 | “I’m pregnant/breastfeeding so unsure about the effects on my child”<br>“I have auto immune disease”<br>“I have a chronic condition/treatment/operation so unsure about effects of the vaccine will have on me” |
|  | Lack of trust in the<br>manufacturer/government/scientists etc. | 9 | “It is not in Government or manufacturers’ interests to tell the truth about side effects and adverse reactions”<br>“The poor management of the pandemic by the government reduces my confidence in the safety and efficacy of a vaccination programme”<br>“Don’t trust it/an American vaccine” |
| **Complacency** | Believe they are not at high risk of COVID-19 | 7 | “I’m not in a risk category”<br>“I don’t want it at this stage as I’m not at high risk of getting COVID” |
|  | Believe they are in good health / Their body can fight off the virus | 6 | “I prefer my body to deal with it in its own way”<br>“I believe maintaining strong immune system is best defence”<br>“I am not in a risk category and I limit my vaccinations to things that potentially have very serious consequences for me” |
|  | Have already had COVID-19 | 3 | “I’ve had COVID already so should be okay for a few months at least”<br>“Would like to know more about antibodies and the likelihood of getting COVID twice”<br>“I’d want to know if I have the antibodies already” |
| **Convenience** | Other people need it more | 7 | “More at-risk people need it first”<br>“It should be delivered to needy first, I’ll have to wait for offer”<br>“I’m fit and healthy that there are more vulnerable people who need it before I do” |
|  | Lack of knowledge about the vaccine | 18 | “I would like to know more about it”<br>“I need to be educated about it first”<br>“I want more information and I need to research about it before accepting it” |
|  | Don’t like injections/vaccine experience | 2 | “I have been told it is very uncomfortable”<br>“The fear of the injection. I have always avoided them” |
|  | Inconvenience | 1 | “Inconvenient” |
|  | Freedom of choice | 1 | “If it were a requirement by law, I would not want it, freedom of choice is important” |

**Table 1b:** Common reasons for vaccine acceptance: survey findings

| Themes | Count | Example of responses |
| --- | --- | --- |
| **Self-protection** | 208 | “To protect me from getting COVID-19”<br>“I’m in a vulnerable group”<br>“It would make me feel safer” |
| **Protect specific others (e.g., family, friends, colleagues etc.)** | 57 | “I want myself, my loved ones, and my community to be safe”<br>“Don’t want to catch the virus and give it to my family”<br>“Want to protect myself and my family” |
| **Protect the population/non-specific others and control the virus** | 139 | “Vaccines are important not just to protect ourselves but others and essential to stop the spread” |
|  |  | “To protect the vulnerable who can’t take the vaccine”<br>“It may save many lives”<br>“The need for herd immunity via vaccine is very important and there needs to be a critical mass of people taking this up” |
| **Confidence in SARS-Cov-2 vaccine** | 87 | “It has been clinically tested and I trust the process”<br>“I don’t believe a vaccine once approved would be unsafe”<br>“It has shown to be effective” |
| **Hope to end the pandemic/ wish for normal life** | 185 | “I want to be able to resume my life”<br>“So that life can get back to normal” |
|  |  | “I just want to be able to hug my daughters”<br>“Truly get on top of this virus and get all our lives and the economy and health service back in action” |
| **Civil duty/Requirement** | 21 | “Everyone who can, should have it. Vaccines are our best chance of eradicating it”<br>“It’s my social responsibility”<br>“I would feel it was my duty, to help to protect other people” |
| **Non-specific pro-vaccine/pro-science statement** | 49 | “I believe in science”<br>“Vaccine works” |
|  |  | “I would take any vaccine at this point”<br>“Can’t think of a good reason why not to take it” |

### 1.2 Rapid Systematic Literature Review

To identify additional themes/reasons for COVID-19 vaccine hesitancy that may have not been captured in our survey, a rapid systematic literature review was conducted. Four electronic databases (Medline, PsychInfo, Medrxiv, PsyAxiv) were searched to identify peer-reviewed journal articles and pre-prints which examined reasons for COVID-19 vaccine hesitancy dated between 01/01/2020 and 03/12/2020: using the following search terms: (COVID-19 vaccine hesitancy) OR ((COVID-19) AND (Vaccine hesitancy)). One researcher (RJ) conducted abstract and full-text screening to determine eligibility and a second cross-checked all eligibility decisions (KA).

Following title and abstract screening, 49 articles remained for full-text screening, with 10 ultimately deemed suitable for inclusion summarised in table 2 [15–24]. The primary reason for excluding articles at the full-text screening stage were that many studies looked at vaccine intention only, not reasons for vaccine hesitancy (see Figure 1). Three of the studies were conducted in the United States, and two in the United Kingdom. The remaining 5 studies were conducted in Nigeria, Mainland China, Hong-Kong, France and Malta respectively. Six studies identified reasons for vaccine hesitancy based on survey questions where a pre-selected list of potential reasons were given. Three studies coded free-text responses to survey questions and one study analysed participant interviews. Six of the studies collected data from general population sample, three collected data from healthcare workers and one did both.

**Figure 1.**
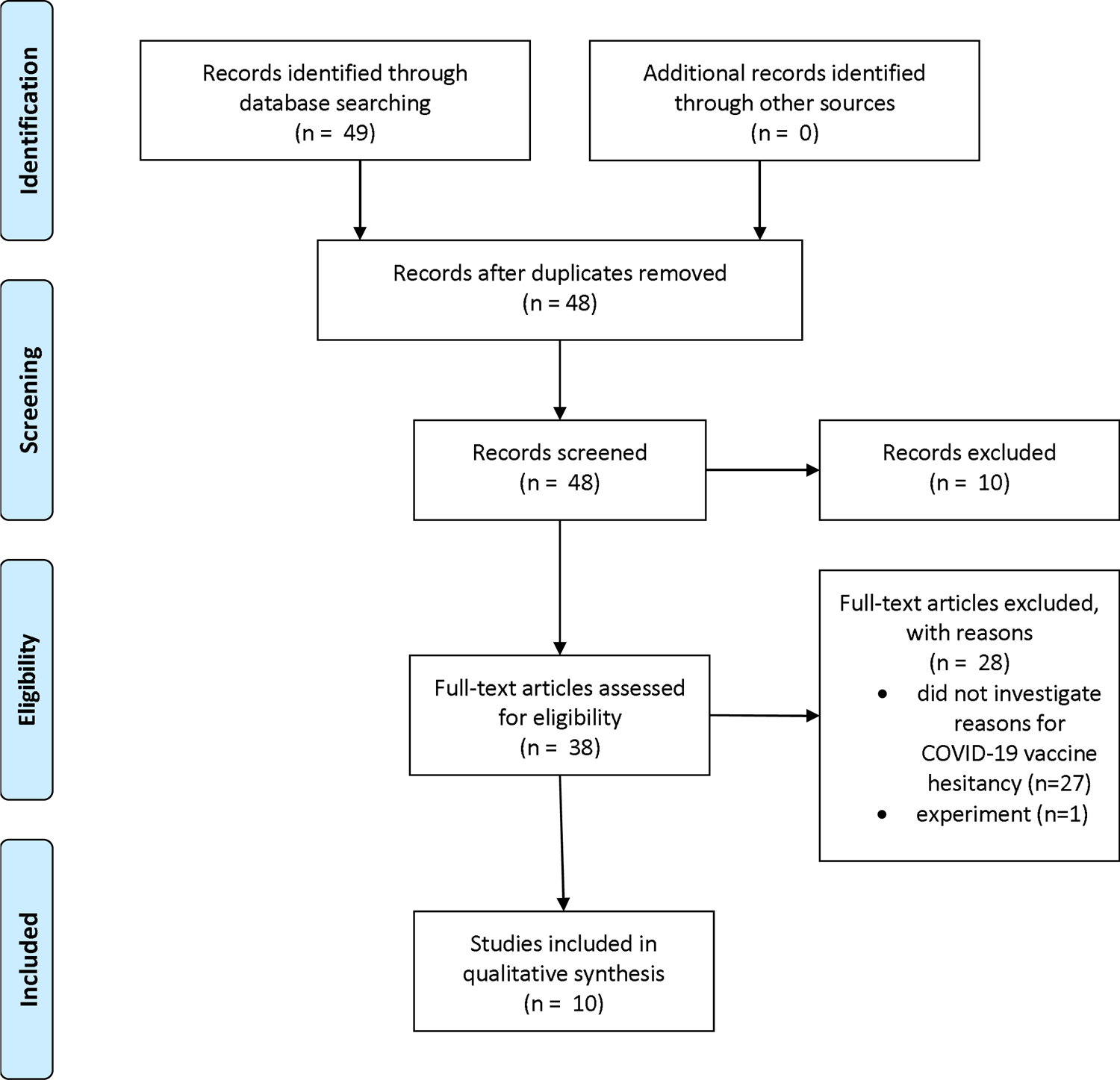
PRISMA summary of search procedure

**Table 2.** Summary of studies included in rapid literature review

| Author | Region | Study design | Population | Sample size | Themes or responses with frequencies<sup>a</sup> |
| --- | --- | --- | --- | --- | --- |
| Adebisi et al., 2020 | Nigeria | Survey question with listed answers | General public | N=517 (n=132 provided reasons for vaccine hesitancy) | Unreliability of the clinical trials (37.1%); immune system is sufficient (27.3%); the vaccine is not safe (16.7%); COVID-19 vaccine is likely to be expensive (6.8%); other reasons (12.1%) |
| Fisher et al., 2020 | US | Open ended question | General public | N=1003 (n=303 provided reasons for vaccine hesitancy) | Specific concerns about the vaccine (82.6%, side effects/safety, efficacy, newness including not wanting to be the first to get the vaccine, rigor of testing, vaccine contents).<br>Need additional information (24.7%, compatibility with personal health conditions e.g., allergies, comorbid conditions, recommendation from doctor or official, timing regarding state of pandemic, personal immunity, need more information unspecified). |
|  |  |  |  |  | Anti-vaccine attitudes, beliefs, and emotions (76.6%, don't need the vaccine e.g., not at risk, religious beliefs, don't believe the vaccine will work informed by reference to other bad vaccine experiences/flu shots not working/vaccine won't work against mutation organism, general statements about not getting vaccines, not comfortable with vaccines, fear about vaccines, misconceptions/incorrect information about vaccines).<br>Lack of trust in vaccines, government and Centers for Disease Control and Prevention (CDC), pharmaceutical companies, vaccine development or testing process, reference to specific conspiracy theories, distrust unspecified (45.2%).<br>Other (9.8%, altruism i.e., wanting higher risk individuals to get first, cost, dislike of needles). |
| Fu et al., 2020 | Mainland China | Survey question with | Healthcare workers and | N= 541 (n=445 provided | Concerns about vaccine safety: newness of vaccine, effectiveness of the vaccine. Cost of the vaccine |
|  |  | listed answers | general population | responses in relation to vaccine hesitancy) |  |
| Gadoth et al., 2020 | US | Free-text question | Healthcare workers | N=1069 (n=609 provided responses in relation to vaccine hesitancy) | “I’m confident there will be other effective treatments soon” (1%)<br>“I don’t yet know enough about the vaccine to make a decision” (14%)<br>“I want to gain natural immunity to the virus that causes covid-19” (2%)<br>“Development of the vaccine may be rushed/the vaccine may not be thoroughly tested prior to approval” (15%)<br>“I believe vaccines may give you the disease they are designed to protect against” (1%)<br>“I don’t know” (1%) |
| Grech et al., 2020 | Malta | Survey question with | Family physicians | N=350 (n=123 provided | The majority of the COVID-19 vaccine related concerns were long term side effects and insufficient knowledge about the vaccine. |
|  |  | listed answers | and trainees | responses in relation to vaccine hesitancy) | Other concerns included: short term side effects (e.g., fever), vaccine effectiveness and general anti-vaccine attitudes. |
| Hacquin et al., 2020 | France | Interviews | General public | N=5028<br>(n=1004 provided responses in relation to vaccine hesitancy) | General opposition to vaccines; concerns that the vaccine would not be effective; not personally required (don't need to get vaccinated); lack of trust in government and pharmaceutical industries. |
| Kwok et al., 2020 | Hong Kong | Survey question with listed answers from a scale | Nurses | N=1205<br>(n=1205 provided responses in relation to | Confidence in safety; effectiveness; and trust in other authorities.<br>Complacency regarding whether the disease is common; that the immune system is sufficient to fight off the disease and the disease is not severe.<br>Constraints to getting vaccinated such as everyday stress; |
|  |  |  |  | vaccine<br>hesitancy) | inconvenience; visiting the doctors; discomfort.<br>Calculations involving weighing up benefits and risks; needing to closely consider whether it is personally useful; needing to understand more about vaccines and vaccination.<br>Collective responsibility including, it not being necessary to get the vaccine when everyone is vaccinated; getting vaccinated can enable an individual to protect people with weaker immune systems; vaccination is a collective action to prevent the spread of diseases. |
| Pogue et al., 2020 | US | Survey question with listed answers | General public | N=316 (33.5% provided responses in relation to vaccine hesitancy) | Concerns about vaccine safety (45.5%); lack of trust in the source that encouraged them to receive the vaccine (13.5%); other e.g., need more testing on the vaccines (15.5%). |
| Sherman<br>et al.,<br>2020 | UK | Survey<br>question with<br>listed answers<br>from a scale | General<br>public | N=1500<br>(n=1448<br>provided<br>responses in<br>relation to<br>vaccine<br>hesitancy) | Concerns about safety and side effects of the vaccine; newness of the vaccine; needing sufficient information to make an informed decision; afraid of needles; not at risk of serious illness from covid; trust in manufacturers/government/health care professionals; |
| Williams<br>et al.,<br>2020 | UK | Free text<br>question | General<br>public | N=527 (n=158<br>provided<br>reasons for<br>vaccine<br>hesitancy) | Concerns about vaccine safety (100%) centred on the newness of the vaccine and its safety (e.g., long-term effect, side effects) and effectiveness. |

<sup>a</sup>Themes or responses were based on participants who provided information on vaccine hesitancy.

The most common themes identified in this review mirrored those identified in our survey. However, the following additional themes were identified: (1) vaccine scepticism; (2) cost of vaccines; (3) concerns relating to vaccine contents; (4) timing of vaccination in relation to the state of the pandemic and (5) concern that the vaccine might result in COVID-19 disease (see table 2).

### 1.3 Additional insights from Public and Patient Involvement (PPI)

To ensure we captured the views of ethnic minority groups on COVID-19 vaccines, we also consulted PPI findings available through the University Hospital Southampton NHS Foundation Trust PPI team regarding the acceptability of vaccines. Several PPI meetings were held on this broad area between July-October 2020, including meetings that specifically sought the views of Black and Minority Ethnic (BAME) individuals.

The feedback from all the consultation meetings was reviewed and was found to reveal considerable overlap in the vaccine concerns identified in these meetings, with those identified as part of our survey and literature review. The only additional concerns related to whether vaccines had been tested on people from different ethnic groups and issues of trust in the medical and scientific communities. These issues were, therefore, prioritised for inclusion in our intervention.

### 1.4 Synthesising findings from Stage 1 to identify most common reasons for vaccine <u>hesitancy</u>

The evidence emerging from the survey, rapid literature review and PPI findings was then triangulated through discussion between the behavioural scientists contributing to this stage of the work. The aim of these discussions was to identify the most common COVID-19 vaccine concerns. This was based in part on the frequency with which concerns were identified in the survey, review, and PPI findings; ensuring that all three domains of the WHO 3C model were represented and that any unique perspectives raised by ethnic minority participants were also captured.

This led to the identification of nine core COVID-19 vaccine concerns. In keeping with the most frequently cited concerns being related to ‘confidence’, 5/9 concerns related to ‘confidence’ (i.e., generalisability of evidence on vaccine safety and effectiveness to diverse populations; side-effects; rapid nature of vaccine development; clinical effectiveness and vaccine scepticism). Two out of ten concerns related to ‘complacency’ (i.e., low perceived risk of COVID-19 and belief in ability to fight off the infection naturally). A further two concerns related to ‘convenience’ (i.e., perceived lack of knowledge about COVID-19 vaccine and altruistic beliefs regarding others having a greater need). A tenth concern was subsequently added when the UK government decided to alter the dosing schedule from 3/4 weeks to up to 12 weeks between the two doses recommended for the Astra Zeneca and Pfizer vaccines. In keeping with the WHO 3C model, this latter issue also related to the issue of ‘confidence’.

### Stage 2: Synthesising the evidence-based views of independent experts

Following the identification of 10 core vaccine concerns (Table 3) we sought to gather evidence-based responses to these concerns. This was achieved through semi-structured interviews with six academic and clinical experts from the fields of public health, general medicine, respiratory medicine and immunology with particular expertise in COVID-19 and/or COVID-19 vaccines. Each expert was presented with the list of 10 concerns and asked to provide an evidence-based response to each concern based on their knowledge of the scientific literature at that time. Interviews with experts were subjected to rapid thematic and content analysis after each interview and interviews continued until saturation in responses was achieved (i.e., no new responses emerged) [25].

**Table 3:** Expert responses to 10 most common reasons for vaccine hesitancy

| Concern | Key responses |
| --- | --- |
| *“I don’t know if the vaccines have been tested on people like me:*<br>*• By age, ethnicity, and comorbid health condition”* | • The vaccines have been trialled in 10s of 1000s of people across many countries and ethnicities<br>• No discernible difference in response to the vaccine across ethnic groups or age groups<br>• Researchers included individuals with common chronic health conditions in the trials to ensure any risks to this population were identified<br>• Pregnant and breastfeeding women were not included in the trials |
| *“I don’t think we know enough about the side-effects of the vaccines”* | • All COVID-19 vaccines have undergone very robust testing, including pauses to trials to explore whether adverse events or allergic reactions were as a result of the vaccine itself<br>• These vaccines follow the same trial protocols for reporting adverse events to the medical advisory boards that all other vaccines must follow<br>• All vaccines come with the chance of immediate side effects, such as a sore arm, fever etc. This shows the immune system has responded to the vaccine<br>• Short term side effects are similar to all other vaccines |
|  | • Although there is less safety data available, MRNA vaccines have been studied for years |
| *“I think the whole process has been rushed”* | • The vaccines have followed the same development criteria that all vaccines must undergo<br>• Many other vaccines are developed in a similar time frame, such as the flu vaccine.<br>• The difference in timeframes has resulted in the concerted channelling of funds into the development of these vaccines, with governments, manufacturers, and scientific bodies providing substantial and rapid funding, expediting the researchers’ ability to test the vaccines<br>• Some vaccines, such as the Oxford Astra-Zeneca vaccine, were developed quickly because the researchers utilised an existing vaccine formula and inserted in an inert form of the COVID-19 virus.<br>• New technology also allowed us to identify the genetic make-up of the virus much more quickly<br>• Evaluation of the safety of the vaccine by independent regulators (MRHA) was expedited as the regulators prioritised reviewing the trial data |
| *“I don’t know if they will work”* | • The data suggests short-term protection of at least 3 months<br>• Pfizer vaccines is highly effective in the short term – |
|  | approximately 95%<br>• Oxford-AstraZeneca rates varied, but were approximately 70% effective<br>• However, long-term data has yet to be reported<br>• We don't know yet if the vaccines prevent transmission |
| *"I don't think I am at risk of getting COVID-19"* | • Whilst many people experience mild symptoms, COVID-19 is unpredictable; we are not able to predict who will be adversely affected.<br>• Although COVID-19 affects older people most severely, a significant proportion of those hospitalised are under the age of 60.<br>• We know that you can contract COVID-19 more than once and are unsure how long any immunity to the virus lasts after exposure.<br>• The vaccines offer protection against the virus and prevent the risk of experiencing a severe form of the disease.<br>• Receiving a vaccine could prevent you from requiring hospitalisation.<br>• Vaccination reduces the volume of the population who can contract and spread the virus, reducing the disease burden in the community. |
| *"I think my body can fight"* | • Younger individuals are less likely to experience |
| *the virus on its own”* | severe COVID-19, however there is still the risk of this happening.<br>• It is also possible to get re-infected with the virus, although evidence suggests the reinfection results in less severe illness.<br>• The immune system can exhibit extreme reactions to the COVID-19 virus, but it is very unlikely to react in such a way to the vaccines.<br>• Reducing your risk of contracting and therefore spreading COVID-19 helps to protect others.<br>• Reducing your risk of contracting COVID-19 also means you are much less likely to need to self-isolate. |
| *“I just don’t know enough about it:*<br>*Safety and effectiveness concerns”* | • The vaccines all significantly reduce the risk of contracting severe COVID-19.<br>• Effectiveness has been shown in individuals of all ages, ethnic backgrounds, and with other health conditions.<br>• No serious side effects have been reported; participants in the early trials have now been monitored for almost 12 months.<br>• The MHRA have been monitoring the vaccines’ safety extremely carefully, as they do with all other vaccines. |
| *“Other people need it more than me”* | • The Joint Committee for Vaccines and Immunisations (JCVI) has identified a priority list for vaccine dissemination.<br>• If someone is offered a vaccine, it means they have been identified as being in a priority group.<br>• Receiving a vaccine does not detract from someone else receiving a vaccine. |
| *“I don’t believe in vaccines: Safety and effectiveness concerns”* | • Vaccines save millions of lives every year and there is no evidence for adverse effects of the COVID-19 vaccines. |
| *“I’m worried I would have to wait 12 weeks before I get my second dose”* | • This decision was taken because it allows twice as many people to get some protection against the virus, offering the greatest opportunity to save lives.<br>• The first vaccination offers short term protection, whilst the second booster dose provides longer term protection.<br>• Delaying the second dose from 3 to 12 weeks also gives the immune system longer to develop immunity.<br>• In the Oxford-AstraZeneca vaccine trials, a longer gap between doses offered better protection. |

The expert responses demonstrated significant thematic overlap and consistency. Table 3 summarises the areas of evidence cited by experts in response to each concern.

### Stage 3: Developing therapeutic dialogues to address common vaccine hesitancy concerns

Our approach to developing the intervention was predicated on two main observations of the existing evidence. First that psychoeducation alone (i.e. provision of information gathered in Stage 2) is unlikely to be an effective way to address COVID-19 vaccine concerns. Second that a central pillar of our approach should be to acknowledge and engage with individuals’ concerns in a supportive context. To achieve this, we sought to develop ‘therapeutic dialogues’ based on the communication principles of motivational interviewing (MI) including:

- *Expressing empathy:* cultivating an empathic space with which to explore hesitancy
- *Developing discrepancy:* identifying areas in which a person’s actions are misaligned with their personal values and goals
- *Embracing resistance:* working collaboratively with an individual to foster change and recognising when that resistance and motivation are intricately tied
- *Supporting self-efficacy:* enhancing confidence that an individual can embark on change[26].

MI was considered an appropriate approach because individuals who are vaccine hesitant are, by definition, not ready to, or ambivalent about, changing their cognitions and behaviour and MI is known to be effective in such contexts.[27, 28] Thus, for each of the most common vaccine concerns identified in Stage 1 we developed a therapeutic dialogue which would both impart information relevant to the individual concern, but do so using the communication principles of MI with a view to facilitating cognitive and, in turn, behaviour change i.e., reduce hesitancy and improve vaccine uptake.

Development of the therapeutic dialogues occurred through several expert workshops with behavioural scientists with expertise in MI, therapeutic interventions, digital interventions, behaviour change and COVID-19. First, key themes identified in the expert interviews (Stage 2) were discussed and translated into conversational language. The investigators chose a conversational approach to align with the online delivery format and to ensure inclusivity for all reading/English levels (see stage 4 below). Second, the dialogues were reviewed to identify points at which MI techniques could be integrated throughout. This process drew on contributors’ experience in behaviour change research and adopted the approach proposed by Rollnick and colleagues [26]. This included *expressing empathy* through use of accepting and non-judgemental language. *Developing discrepancy* by simultaneously providing information related to the concern, and presenting a rationale for vaccine uptake. The latter were derived from survey respondents willing to accept a COVID-19 vaccine (see Table 1b) and sought to develop a discrepancy between the individual’s cause for concern and their wider personal values and goals. *Embracing resistance* by acknowledging that their concerns are shared by others and are legitimate and *supporting self-efficacy* by reinforcing the individual’s personal agency in making their decision to accept a vaccine or not. See Table 4 for illustrative examples of how MI principles were embedded within the therapeutic dialogues.

**Table 4.** Exemplars of how MI principles were included within the therapeutic dialogues

| Concern | Motivational interviewing concept | Concept example utilised in the dialogue |
| --- | --- | --- |
| *"I don't know if the vaccines have been tested on people like me"* | Expressing empathy:<br>Including reflective listening to concerns and integration of follow up questions to engage user | These are brand new vaccines and it is completely understandable that you would ask about their safety |
| *"I don't believe I am at risk"* | Developing discrepancy: | So when you choose to have |
| *of getting COVID-19”* | Identifying potential areas of conflict between vaccine hesitancy and personal values | a vaccination you are also choosing to protect others, to take the pressure off the NHS, and helping us all get back to normal. |
| *“I don’t think we know enough about the side-effects of the vaccines”* | Embracing resistance:<br>Recognising resistance and helping to move forward collaboratively | And you are not alone in wondering about this. Scientists, doctors, the independent regulator who decide on which medicines can be offered to the public (the Medicines and Healthcare Products Regulatory Agency) all want to know how well the vaccines work. |
| *“I don’t know if the vaccines have been tested on people like me”* | Supporting self-efficacy<br>Enhancing confidence to make an informed decision about whether to receive a vaccine | We hope we have been able to help with your concerns about the safety of the vaccines. To sum up, they have all been monitored very closely to find side effects. But if you did experience a side effect it is most likely to be very minor |
|  |  | and much less severe than catching COVID-19. |

Finally, we hosted a PPI workshop to discuss the resulting dialogues. The group, while small (n=4), included two adults less than 30 years (two greater than 50 years); three women and one man and all reported an interest in vaccine hesitancy and had some experiences of it in friends and family. The feedback obtained through this workshop fostered changes to their readability, along with expansion of the information conveyed and greater consideration of specific groups within the population (i.e. those who have allergies or specific religious and cultural needs). No additional vaccine concerns were identified by the group.

### Stage 4: The digital intervention

The script from each of the 10 therapeutic dialogues provided the architecture for our digital, web-based, vaccine hesitancy intervention. This intervention has been built in the form of a conversational interface through which individuals identify the issue that most closely underpins their reason for being hesitant, (from the issues stated above e.g., concerns about side effects). This identification triggers a therapeutic dialogue relevant to the selected concern, with opportunities for the individual to further explore the content as they progress through the dialogue as well as to access responses to more than just their initial concern.

Once developed, the digital intervention was piloted with 18 members of the public (nine male / nine female) who had no previous experience with the dialogues. Participant feedback on the user interface, accessibility, and general presentation led to a final iteration of the intervention, which can be viewed here www.covidvaxfacts.info1

### Discussion

The development of safe and effective vaccines against SARS CoV-2, while necessary, will not be sufficient to contain COVID-19 unless we also achieve high vaccine uptake. We have described here the rapid development of an evidence-based digital intervention which draws on the communication principles of MI and is in keeping with many of the recommendations made in a recent review of approaches to increasing vaccine uptake e.g., focus on the concerns of the population.[29] Our aim is to provide the end-user with an intervention which is individualised to their specific concerns, acknowledges the legitimacy of these concerns, provides up to date information related to these concerns whilst also providing an accepting non-judgemental context in which they can explore their reasons for hesitancy. The text-based content and digital format mean it can be readily scaled-up for wider dissemination and rapidly modified for implementation in different languages and to respond to changing information.

Although this intervention, like much else to do with COVID-19, has been developed at pace we think the process highlights some potential issues regarding intervention development worthy of discussion. First, the development of our digital, behavioural intervention followed a fairly conventional path as outlined in the Medical Research Council’s (MRC) best practice guidance. This involved evaluating the evidence base and theory as well as incorporating the views of target users [30]. This was possible partly because we had timely access to PPI findings available through the University Hospital Southampton NHS Foundation Trust regarding the acceptability of vaccines, allowing rapid comparison of the PPI findings with the concerns identified through our existing survey data and literature review.

A critical step in digital intervention development is optimisation of intervention content, since digital intervention content cannot be adjusted ‘in the moment’, like in a practitioner delivered intervention. We were able to conduct optimisation work with PPI, albeit with a smaller sample (N=4) than might usually be employed in digital intervention development. Computer science methodology states that during intervention optimisation around 80% of views can be captured with five target users and we were close to this threshold [31].

However, best practice guidance from digital health psychology suggests including larger, diverse samples is important to ensure views of people from different backgrounds are considered [30]. Despite having a smaller sample, our optimisation with PPI did help us to improve the persuasiveness and accessibility of the key messages within the intervention. It is possible that we may have found other important ways of optimising our content by including a larger, more diverse group of PPI at this stage. However, it is important to note that this intervention is quite simple, it targets only one behaviour, draws on a very well-established behavioural technique which guided content design (MI), and it addressed barriers that were thoroughly identified using existing evidence in the intervention planning stage. Therefore, in this particular context, it is possible that sufficient optimisation was achieved with a smaller sample.

The MRC highlights the importance of making use of existing data and evidence wherever possible. In this work, we were able to benefit from data collected as part of another study[14] where we were able to identify specific concerns related to vaccine hesitancy. We also drew on evidence kindly shared with us by others. This allowed acceleration of the intervention development and improved the economic efficiency of research.

In view of the urgency of the public health issue we conducted a rapid review. Indeed, COVID-19 has most likely led to an unprecedented number of rapid reviews, as the scientific community have clamoured to understand the available evidence as quickly as possible.

Although, it is clear that rapid reviews take many forms (e.g., limited by language, dates, databases etc.), they do vary in the quality of their reporting and the methodological shortcuts they take.[32] The implications of these inconsistencies for the quality and validity of these reviews is, however, unclear as there is thus far limited evidence comparing the results of different review approaches. The provision of such evidence in future research would undoubtedly inform the contexts in which it is appropriate to conduct rapid reviews, and the methods that should be employed. Such guidance now exists for scoping reviews[33] and would appear to be in development for rapid reviews by the Equator network.[34]

### Conclusion

In summary, for COVID-19 vaccines to achieve their full public health potential, the public need to be willing to be vaccinated. Recent data suggest this cannot be assumed. We have reported here on the development of a scalable digital intervention which seeks to address the concerns of individuals who are vaccine hesitant with a view to enhancing their confidence in COVID-19 vaccines and, in turn their uptake. The effects of the intervention on these outcomes will be the subject of future work.

### Footnotes

^1^Web URL is not currently active but will be active by the time of publication

### Funding

This work was supported by the School of Primary Care Research [Grant Number 434].

### Authorship contribution statement

HK, KV, KB, KB, TC: conceptualisation, data analysis, writing original draft, reviewing and editing. RJ, KA: Conducted rapid literature review, writing of original draft, reviewing and editing. WSL, JRM, LD: conceptualisation, expert contributions through qualitative interviews, reviewing and editing. TA, JB, CB, RB, TMK: reviewing and editing.

## Data Availability

Literature review data are available upon request.

## Acknowledgements

We would like to thank Dr Lucy Fairclough (University of Nottingham), Dr Simon Royal (University of Nottingham), and Dr David Turner (University of Nottingham) for their contributions to the expert interviews.

